# Cortical Visual Processing Differences in Myopia and Blur

**DOI:** 10.1101/2024.03.22.24304534

**Authors:** Katia Steinfeld, Micah M. Murray

**Author notes:** The authors declare no competing financial interests.

## Abstract

Myopia is projected to impact over 50% of the global population by 2050. Despite links to long-lasting anatomical changes in visual cortices, little is known of potential consequences of myopia on visual brain functions, such as visual completion. We hypothesized that adults suffering from moderate myopia process attentionally demanding visual stimuli under optical blur differently than emmetropic adults. Non myopes (*N*=12) and low-to-mild myopes *(N*=13) were tested under −3 diopters of lens-induced blur. Participants performed an illusory contour discrimination task while 128-channel EEG was recorded. Each trial also included an intervening, task-irrelevant dartboard stimulus. Visual evoked potentials (VEPs) to the illusory contour (IC), no contour (NC), and dartboard stimuli were analyzed using an electrical neuroimaging framework. We provide evidence for cortical processing differences between non myopes and mild myopes at 218-2800ms post-stimulus during visual completion, but not during viewing of dartboards. These differences stemmed from topographic modulations, indicative of the engagement of distinct networks of brain regions that were localized to medial portions of the occipital pole. Moreover, the predominant VEP topography during this time period both correlated with extent of refractive error and also was an excellent classifier of myopia vs. emmetropia. By contrast, our analyses provided no evidence for differences in visual completion processes between groups. To our knowledge, this is the first study of myopia pairing high-density EEG and a behavioral task. Collectively, this pattern of findings supports a model of myopia wherein low-level visual cortices are impacted at relatively late post-stimulus processing stages.

## Introduction

By 2050, myopia’s prevalence is expected to reach 52% of the global population (Holden *et al*., 2016). For effective prevention and treatment, it is essential to characterize myopia’s downstream chain of events (Wildsoet *et al*., 2019). Defocus and its neural translation (Maiello *et al*., 2017) play a central role in myopia. Myopia onsets when the eyes continue to grow past their focal distance (Wolffsohn *et al*., 2019). It is most frequently diagnosed in late childhood and progresses (Chua *et al*., 2016) during developmental states when neural pathways remain particularly plastic (Atkinson and Braddick, 2013). In myopia, defocus and eye growth are interrelated. Animal models have shown that induced retinal blur triggers eye elongation (Chakraborty *et al*., 2020). Furthermore, in both corrected myopic adults and children, there is significant impairment in paracentral retinal processing (Chen *et al*., 2006; Ho *et al*., 2012; Gupta *et al.,* 2022), the magnitude of which correlates with myopia’s progression (Luu *et al*., 2007). Retinal blur is thus part of a feedforward loop of eye elongation, which results in a lifetime impairment in retinal biometric parameters (Zhang *et al*., 2023) and foveal responses (Li *et al*., 2017; Haarman *et al*., 2020).

There is also nascent evidence for lifetime changes in the brains of myopic patients, both with low (LM) and high myopia (HM), i.e. below and above −6D, respectively (Troilo *et al*., 2019). Cheng *et al*., (2020a) describe a significant decrease in LM subjects’ resting-state activity in V1 and the optic radiations, when compared to HM patients and emmetropes. The same group documented decreased connectivity within higher-order visual cortices in the right para-hippocampal gyrus of LM and HM individuals versus emmetropes (Cheng *et al*., 2020b). The collective implication is that myopia may impact both lower-order and higher-order visual regions.

To our knowledge, the cortical consequences of myopia have never been investigated with high-density EEG methods nor an electrical neuroimaging analysis pipeline. The technique provides (sub) millisecond temporal resolution, allowing for precise characterization of both the amplitude and the topography of cortical activity over time during a visual task (Michel and Murray, 2012). These robust EEG analyses have been applied to characterize sensory processing (Matusz *et al.,* 2015; Retsa *et al*., 2020; Thelen *et al*., 2014; Turoman *et al., 2021*) and pathologies (Berchio *et al.,* 2023; Biria *et al.,* 2018; Jan *et al*., 2019; Layer *et al*., 2022, Retsa *et al*., 2023).

Additionally, myopia and its defocus alter visual experiences during years when children are still fine-tuning their visual system. Illusory contour (IC) sensitivity, for one, matures through adolescence (Hadad *et al*., 2010; Altschuler *et al*., 2014). These processes allow an individual to perceive fragmented elements as continuous surfaces and unobstructed objects (Murray and Herrmann, 2013). During childhood it involves a distributed network over occipital and frontal cortices, in a relatively effortful process (Hadad *et al.,* 2010; Altschuler *et al*., 2014). With visual experience, the processing of such illusions gains in speed and is consolidated across the ventral visual stream associated with object processing (Ringach and Shapley, 1996; Mendola *et al*., 1999; Murray *et al*., 2002). As such, one can postulate that the defocus experienced over a lifetime in adults with myopia may impact perceptual completion processes, including illusory contour sensitivity.

Recent evidence points to long-lasting changes in both retinal and cortical dynamics in myopia. Furthermore, defocus seems to play a central part in myopia’s progression. Here, we characterize the dynamics of the interaction between myopia and optical blur in the cortex. We hypothesized that adults with low and moderate myopia process optical blur differently than emmetropic adults. To test this hypothesis, we used an illusory contour recognition task while acquiring high-density EEG recordings. To our knowledge, there have been no similar brain imaging/mapping studies of visual function that compared myopic participants with corrected-to-normal vision and emmetropic participants (though see McKyton *et al*., 2015 and Hadad *et al.,* 2017 for behavioral studies of children with residual myopia following removal of congenital cataracts). As such, mechanistic differences in the processing of illusory contours remain undefined. Because we hypothesized that optical blur has different effects in myopia’s and emmetropia’s visual processing, all participants were tested under the same level of induced defocus.

## Methods

### Participants

Participants were recruited from among students at the University of Lausanne and Lausanne University Hospital. All participants underwent an orthoptic evaluation, including automated refraction, subjective refraction, and ocular motility testing. Exclusion criteria were history of neurologic, psychiatric, or ophthalmic disease, corrected visual acuity less than 0.00 LogMar in any one eye, or astigmatism equal or superior to −1D in any one eye. Two participants were excluded based on such criteria. One for high astigmatism and the other for having undergone myopia-correction surgery. Additionally, one myopic participant received an updated prescription with which they reached 0.00 LogMar visual acuity, worn during the experiment. Data from twenty-five participants were included in the analyses presented here, 13 myopes (males *n*=4, females *n*=9) and 12 non myopes (males *n*=4, females *n*=9). Because there were no significant differences in spherical equivalent of refractive error between each subject’s eyes and none of the subjects were hyperopic (see Table 1), we refer to the refractive error as the absolute value of the mean between both eyes’ spherical equivalent (sphere + cylinder/2). The Cantonal Ethics Committee approved this project (protocol #2018-00240). All participants provided written informed consent after verbal and written explanation of the study, according to the tenets of the Declaration of Helsinki.

**Table 1.1.**
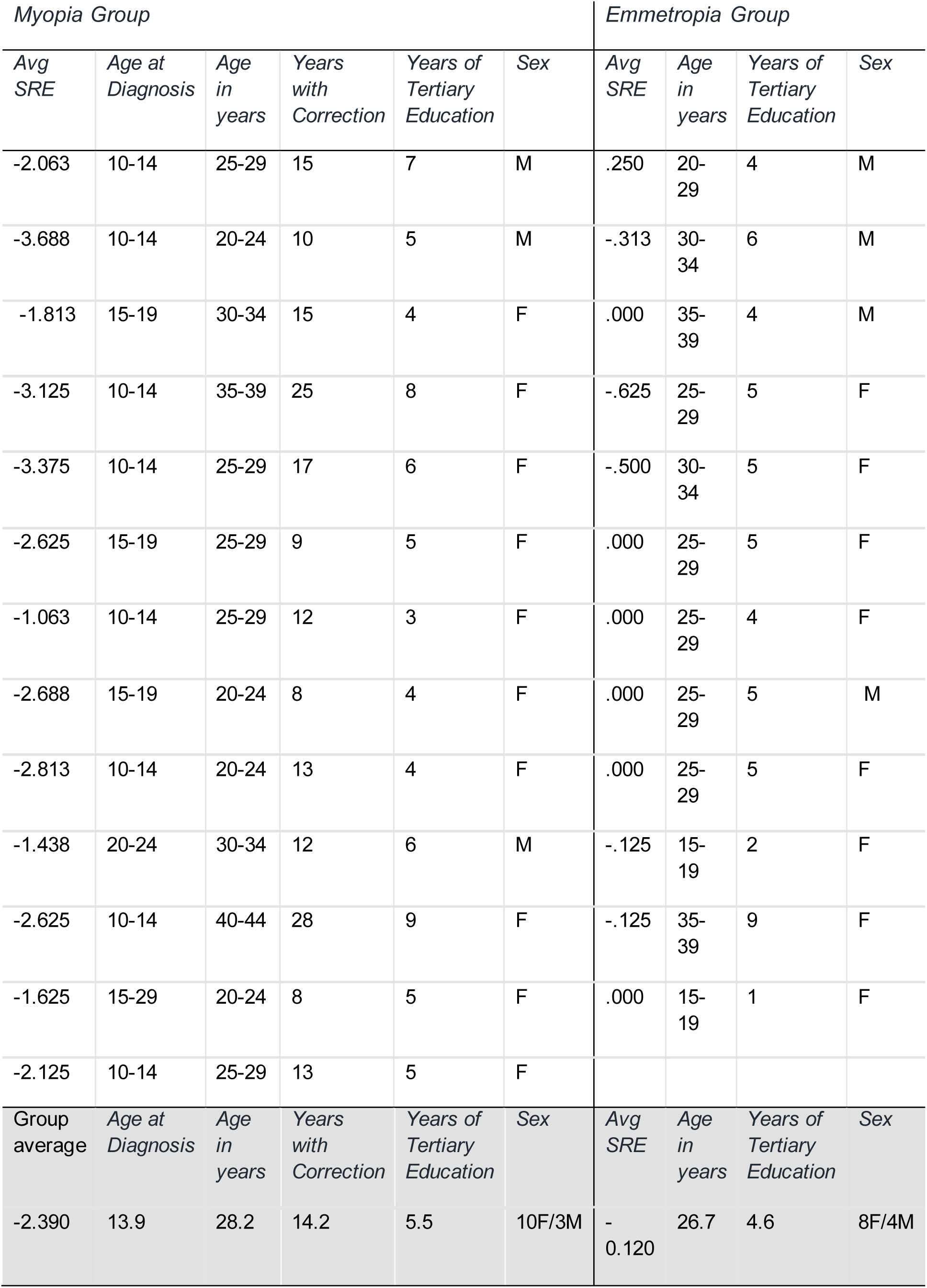
Participant’s average of both eyes’ spherical equivalent, age of diagnosis, age at time of testing, years with correction, years of tertiary (post-secondary school) education, and sex.

### Stimuli and Protocol

Participants sat at 60cm distance from a 20-inch LCD monitor, 1600 x 1200 @ 60Hz, pixel response time 16ms, display 100 PPI. The emmetropia group wore only one frame with +3D convex lenses to simulate –3D myopia. The myopia group wore their corrective lenses as well as the +3D convex lenses. Participants had both eyes un-occluded. During breaks they were allowed to take off the frames and were advised to close their eyes to avoid fatigue. For the experiments, participants viewed five stimuli: a checkered dartboard, two illusory contour shapes (ICs) and two no contour (NC) images, as detailed below.

The IC stimuli were Kanizsa-style squares and circles formed by four ‘pacman’ inducers which appear black on a gray background at 50% contrast. Each pacman’s diameter was 6.96° of visual angle. The IC square (IC_square_) sides subtended 9.52° of visual angle with a support ratio of 50%. Support ratio refers to the percentage of the contour physically present versus the entire contour of the perceived shape, such that a support ratio of 100% would mean that the perceived shape was represented by a physical contrast gradient. IC_square_ inducers were presented on the diagonal axes with their centers at 6.73° of eccentricity from central fixation. The IC circle’s (IC_circle_) diameter subtended 10.76° of visual angle with a support ratio of 62%. IC_circle_ inducers were presented on the cardinal axes with their centers at 5.38° of eccentricity from central fixation. The no-contour (NC) circle (NC_circle_) and square (NC_square_) inducers were at the same position as in their IC counterparts but were rotated 180° outwards. We chose to use two illusory contour shapes formed by inducers at different positions to avoid an attentional bias, where participants would potentially complete the task by selectively attending certain regions of the visual field. Furthermore, other studies by our group have successfully used these same inducers and illusory contour shapes, providing a benchmark for IC sensitivity markers (Knebel and Murray, 2012; Murray *et al*., 2002).

Participants’ task was to indicate the presence versus absence of an illusory contour, regardless of its shape, via a serial response box (see also Murray *et al*., 2002). Subjects were instructed to press a button labeled ‘1’ as quickly and accurately as possible when they saw a shape, either circle or square, and another button label ‘2’ when they did not. In turn, participants were instructed not to respond to the dartboard stimulus. There were no restrictions regarding hand preference. One participant in the myopia group, and two participants in the emmetropia group responded with their left hand.

The checkered dartboard was a circular, black and white, pattern-reversal stimulus. Its Michelson contrast was 100%. It consisted of 16 wedges and 8 concentric circles or a total of 128 checks. The eight concentric circles had eccentricities of 1.60°, 3.32°, 4.83°, 6.44°, 8.05°, 9.64°, 11.24° and 12.84°. Spatial frequencies within wedges were of 0.3 cpd. Spatial frequencies at the border of each ring ranged from 0.8 cpd (innermost) to 0.1 cpd (outermost). Within this eccentricity, spatial frequency range and contrast, resolution is not a constraint. Every presentation lasted 400ms and reversed in spatial phase at 5Hz (10 reversals per second, 100ms for each pattern). VEPs to these stimuli provide an assessment of general group-wise differences to task-irrelevant stimuli.

The complete trial sequence is depicted in **Figure 1**. It consisted of a dartboard presentation for 400ms followed by a blank interval of 1s on average (random duration ranging 0.8-1.2s), a subsequent pacman array presentation for 400ms, and a second blank interval of 2s on average (random duration ranging 1.8-2.2s). Stimulus order of IC and NC conditions within a block was pseudo-randomized. A block of trials lasted 6min on average and included 80 dartboard presentations interleaved with 80 pacman array stimuli (i.e. 40 IC and 40 NC stimuli). Participants underwent one training session prior to recording and ≥8 recorded blocks of trials.

**Figure 1.**
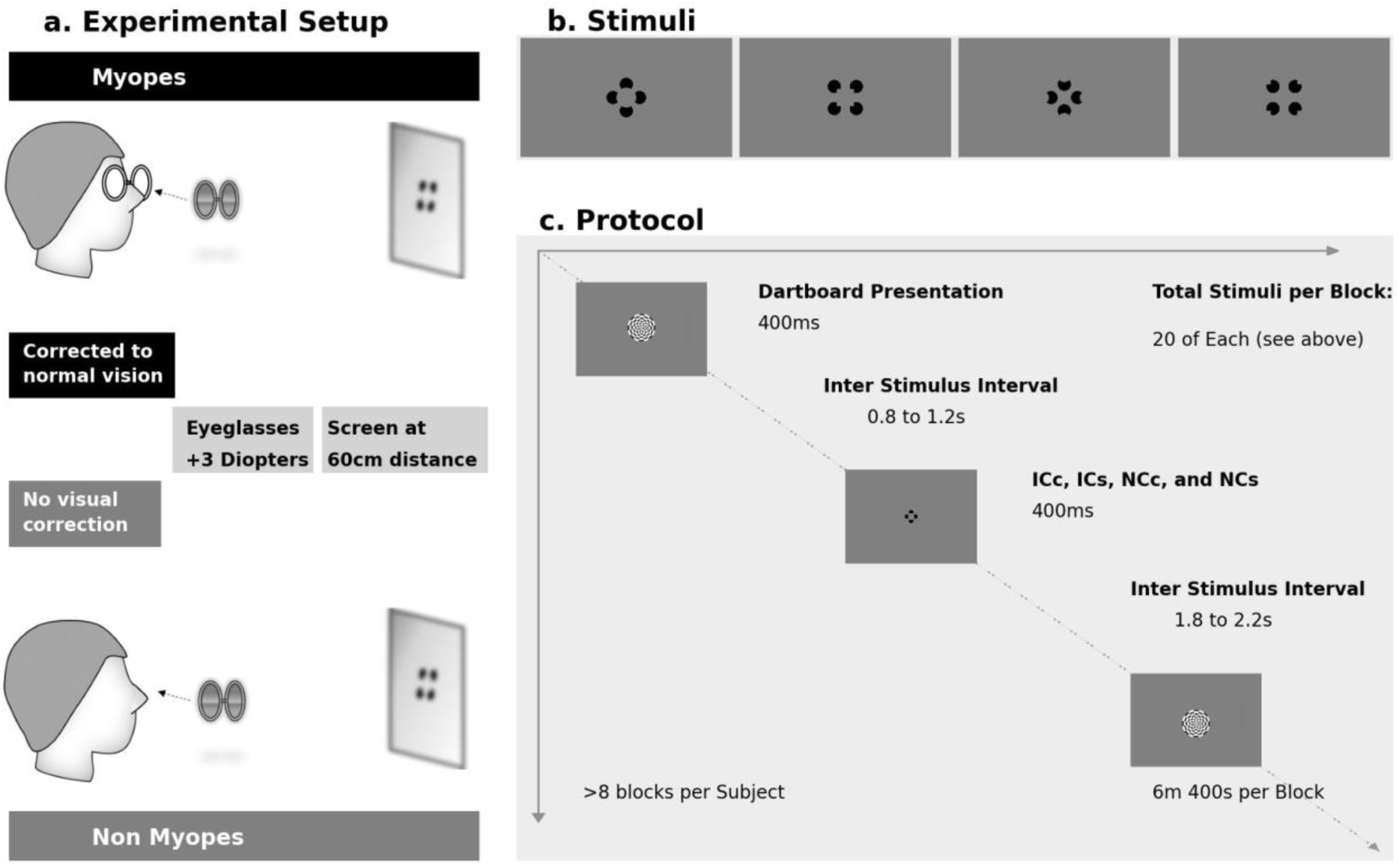
Schematic of the Experiment, Stimuli and Protocol. a. In the experiment, participants viewed stimuli at 60cm distance while EEG was recorded. Myopes wore their usual corrective eyeglasses as well as additional eyeglasses with +3D convex lenses to simulate −3D myopia. Non myopes wore only the +3D convex lenses only. b. Stimuli consisted of illusory contour (IC) circles and squares as well as their no contour (NC) counterparts wherein the pacman inducers were turned outward 180°. Stimulus order within a block of trials was pseudo-randomized. c. A trial sequence entailed the presentation of the dartboard (400ms duration), followed by a 1s blank interval (random duration ranging 0.8-1.2s), an IC or NC stimulus presentation (400ms duration), and an additional blank interval of 2s (random duration ranging 1.8-2.2s).

### Behavioral Analyses

Participants and trials excluded from the analysis according to EEG criteria were also excluded from the behavioral analyses. We exported the behavioral dataset as pandas in Python for calculation of mean reaction times, error rates and false alarm rates. An ‘error’ was computed when participants reported seeing an illusory shape when an NC stimulus was presented. A ‘false alarm’ was counted when participants reported seeing an IC during presentation of the dartboard. False alarm rates were below 1% for all participants and thus they were not examined further. A mixed-model repeated measures ANOVA was performed with the SPSS software (IBM, 2021), with Stimulus (IC vs. NC) as the within-subjects factor and Group (myopia vs. non-myopia) as the between-subjects factor.

### EEG Acquisition and Preprocessing

Continuous EEG was acquired at 1024Hz through a 128-channel Biosemi ActiveTwo AD-box referenced to the common mode sense (CMS; active electrode) and grounded to the driven right leg (DRL; passive electrode). This configuration creates a feedback loop, driving the montage’s average potential towards the amplifier zero. We used the Cartool freeware (Brunet, Murray and Michel, 2011) for pre-processing and analyses. The continuous EEG was filtered with a Butterworth filter (−12dB/octave roll-off, high-pass 0.18Hz, low-pass 60Hz), underwent DC/0Hz removal and were notched at 50Hz. Epochs spanned from 100ms pre-stimulus onset to 500ms post-stimulus onset. All epochs were then tested to the threshold value of ±80μV, in a semi-automated fashion accompanied by visual inspection, to reject epochs with artifacts and transient noise. Epochs were then averaged to create VEPs for each participant and stimulus.

Subsequently, VEPs were then 40Hz low-pass filtered. To identify electrodes that were broken or consistently had bad contact, VEP waveforms and topographies were inspected. Signal from each selected electrode was replaced by an interpolation (Mean±SD= 7.36±4.11 electrodes, max=14, min=0) using three-dimensional splines (Perrin *et al*., 1987). During this pre-processing, 5 participants beyond the 25 mentioned above were excluded due to excessive noise. Following interpolation, data were recalculated using a common average reference. Results presented here are based on data from 13 myopes and 12 non myopes (detailed in Table 1). For each of these remaining participants, 415 or more epochs were accepted in response to the dartboard presentation. There was no significant difference between groups in the number of accepted epochs in response to dartboard presentations (*t(23)*=−0.252, *p*=0.803). Likewise, for all subjects more than 150 epochs were accepted in response to each stimulus condition (i.e. IC_circle_, IC_square_, NC_circle_, and NC_square_; Mean±SD= 597±107 epochs). As our interest here was not in the responses to individual stimulus conditions, we collapsed IC_circle_ and IC_square_ epochs into a single IC VEP for each participant and collapsed NC_circle_ and NC_square_ epochs into a single NC VEP for each participant. There were no significant differences in the final number of accepted epochs per condition (*F*_(1,19)_ =0.2498, *p*=0.128, *η_p_^2^* =0.098) nor between groups (*F*_(1,23)_ =0.654, *p*=0.427, *η_p_^2^* =0.028).

### VEP Analyses

#### Group-wise differences in processing task-irrelevant dartboards

To test for group-wise differences in processing of the task-irrelevant dartboard stimuli we performed a mass univariate analysis of the VEPs. This entailed a series of un-paired t-tests across the full 128-channel electrode montage and as a function of peri-stimulus time. This analysis was conducted with the Cartool freeware (Brunet *et al*., 2011). Effects were considered reliable if they met the following criteria: 1) temporally sustained at a given electrode for ≥10 consecutive samples and 2) present across at least 10% of the 128-channel electrode montage. Similarly, we analyzed the Global Field Power (GFP), which is the root mean square of the voltage of all electrodes and yields larger values for stronger VEPs (Murray *et al*., 2008).The same temporal criterion of 10 consecutive samples was used.

#### Group-wise differences during illusory contour sensitivity

VEP analyses followed a 2×2 mixed model design with the between-subjects factor of Group (myopia vs. emmetrope) and the within-subjects factor of Stimulus (IC vs. NC). For the analysis of voltage waveforms and GFP waveforms, we used the Cartool freeware (Brunet *et al*., 2011) and STEN (Knebel and Notter, 2012). Both the VEP waveforms from the full electrode montage as well as reference-independent measures of the electric field at the scalp were analyzed (Koenig et al., 2011; Michel & Murray, 2012). Statistical effects on VEP waveforms needed to meet both a temporal criterion of ≥10 contiguous time points as well as a spatial criterion of spanning across at least 10% of the electrodes (Guthrie and Buchwald, 1991). Despite the vast number of tests performed, we include these analyses here for visualization purposes and to assist readers who are potentially less familiar with analyses based on multivariate, reference-independent metrics. However, we would emphasize that our interpretations here are based solely on these latter metrics. In terms of multivariate reference-independent measures, we first analyzed the GFP. When analyzing GFP, significant main effects or interactions needed to satisfy a temporal criterion of ≥10 contiguous time points (Guthrie and Buchwald, 1991).

The VEP topography, which is also reference-independent, was analyzed with hierarchical clustering and application of a modified Krzanowski-Lai criterion to select the number and pattern of stable VEP topographies that best characterized the variance in the concatenated dataset (Murray, Brunet and Michel, 2008). This technique maps stable electric field topography, or ‘template maps’, across time for each group and stimulus condition. By visual inspection of the clustering results, periods when ‘template maps’ differed between groups and/or stimulus conditions were determined. All further steps were constricted to each of these time periods, considered separately.

To statistically assess potential topographic differences during these time periods, individual subject’s VEPs were fitted with all the ‘template maps’ identified in these time periods. This fitting procedure entails labelling each time point with the template map with which it best correlates spatially (Murray, Brunet and Michel, 2008). As an output, it yields the total number of time points a given template map fit the data from each subject and stimulus. These values were then submitted to an rmANOVA. This rm-ANOVA had as within-subjects factors of ‘template maps’ as well as Stimulus (IC vs. NC) and the between-subjects factor of Group (myopia vs. emmetropia). We also performed a linear regression between the template maps’ total number of time points with refractive error.

Lastly, source reconstruction was performed for the time periods where clustering results showed a significant difference between Groups and/or Stimuli or their interaction. Firstly, VEPs for each subject were averaged across IC and NC conditions (as only a main effect of Group was observed, as detailed below), cropped to the selected time window and averaged over time. Source modelling was performed with a distributed linear inverse solution (minimum norm) combined with the LAURA (local autoregressive average) regularization approach (Grave de Peralta Menendez *et al*., 2001, 2004; see also Michel *et al*., 2004 for a comparison of inverse solution methods). The solution space was calculated on a realistic head model that included 5923 nodes, selected from a grid equally distributed within the grey matter of the Montreal Neurological Institute’s average brain (available from https://sites.google.com/site/cartoolcommunity/downloads). The head model and lead field matrix were generated with the Spherical Model with Anatomical Constraints (SMAC; Spinelli *et al*., 2000 as implemented in Cartool version 4.10 by Brunet *et al*., 2011) using a 4-shell model (skull, scalp, cerebral spinal fluid, and brain) as well as with an upper skull thickness (5.7mm) and mean skull conductivity (0.021S/m) values based on the mean age of our sample. As an output, LAURA provides current density measures; their scalar values were evaluated at each node. Statistical analysis of source estimations was performed with an un-paired t-test (p<0.05 at a given node) and with a cluster size criterion of minimally 10 contiguous nodes based on randomization thresholds (see also De Lucia *et al*., 2012; Knebel and Murray, 2012; Murray and Herrmann, 2013; Retsa *et al*., 2018, 2020, for similar implementations).

## Results

### Demographics and refraction

The myopia and emmetropia groups did not differ in age (*t*_(23)_=0.68; *p* =0.51; *d*=0.27), sex (*Χ*^2^_(1, 25)_= 0.33; *p*=0.57) or years of education (*t*_(23)_=1.19; *p* =0.97; *d=.*48) (see Table 1). All myopia participants were first diagnosed between the ages of 12 and 21 years, which was ≥8 years prior to this study (range=8-28 years; mean±SD*=*14.23±1.69 years). For all participants, between-eyes differences in sphere (SRE) and cylinder (CRE) refractive error were not clinically relevant (difference in SRE ≤ |1.25|D, in CRE ≤ |0.25|D). In an rm-ANOVA of astigmatism levels where within subjects factor were right and left eyes and between subjects factor were myopia and emmetropia groups, astigmatism was shown to be significantly higher (*F*_(1,23)_=7.246; *p*=0.013; *η_p_^2^*=0.24) in the myopia group (mean±SD in the right eye =-0.462±0.105 and −0.481±0.098 in the left eye) than in the emmetropia group (right eye =-0.083±0.109; left eye =-0.104±0.102). There were no significant differences in astigmatism levels between eyes for either group (*F*_(1,23)_=0.23; *p*=0.63; *η_p_^2^* =.010).

For all 25 participants, we calculated the spherical equivalent from each eye’s sphere and cylinder refractive errors, averaged the spherical equivalents of both eyes and took its absolute value, thus obtaining a unique positive average spherical equivalent value per subject (SRE). Myopia was present in 13 participants (SRE ranged −3.69D to −1.36D; mean±SD = −1.36±1.38D) and absent in 12 participants (SRE ranged −1D to 0D; mean±SD = −0.34±0.28D). For a complete description of myopes’ and non myopes’ refractive errors see Supplementary Table S1 and S2 respectively in supplementary data.

### Behavioral Results

All participants performed at or above 87% accuracy on the illusory contour recognition task. Errors represented >1% of the trials for four participants. When error rates were analyzed with a 2×2 rmANOVA, there was no evidence of a main effect of Group or Stimulus nor any interaction (all p’s >0.34). There was no evidence of reliable differences between groups in terms of accuracy (*F*_(1,23)_=0.010; *p* =0.920; *η_p_^2^* =0.00) or reaction times (*F* _(1,23)_ = 0.48; *p* =0.83; *η_p_^2^*=0.002). There was a main effect of Stimulus on reaction times (*F*_(1,23)_=11.721; *p* =0.002; *η_p_^2^*=0.338), with detection of ICs being significantly faster than that of NCs (mean±SD = 466±17ms vs. 500±17ms). However, accuracy was not significantly different between stimulus conditions (*F*_(1,23)_=0.763; *p=0.391*; *η_p_^2^* =0.032). The interaction term was not significant for either the analysis of accuracy rates (*F*_(1,23)_=1.261; *p*=0.273; *η_p_^2^*=0.052) or reaction times (*F*_(1,23)_=0.678; *p*=0.419, *η_p_^2^*=0.029). To investigate possible speed-accuracy trade-offs, we computed inverse efficiency scores, i.e. mean reaction times divided by percentage of correct trials (Townsend and Ashby, 1978, 1983). The rmANOVA on inverse efficiency scores did not reveal a significant effect of Group (*F*_(1,23)_=0.055; *p*=0.816; *η_p_^2^*=0.002) nor a significant Group × Stimulus interaction (*F*_(1,23)_=1.194; *p*=0.286; *η_p_^2^*=0.049). The significant main effect of Stimulus did, however, persist (*F*_(1,23)_=5.907; *p*=0.023; *η_p_^2^*=0.204), confirming that the speed gain in IC detection did not come at the cost of accuracy.

### Neurophysiological Results

#### Dartboard Stimuli

We analyzed VEPs in response to the task-irrelevant dartboard stimuli. Exemplar VEP waveforms and GFP waveforms are shown in **Figure S1**. A mass univariate analysis (i.e. un-paired t-tests at each electrode as a function of time peri-stimulus), failed to identify temporally sustained significant differences at more than 10% of the electrode montage. We likewise conducted an analysis of the GFP in response to the dartboard stimuli that also failed to identify temporally sustained significant differences. These results provide no indication for group-wise differences in processing of task-irrelevant stimuli.

#### Illusory Contour Sensitivity

In light of our laboratory’s prior works on brain mechanisms of illusory contour sensitivity in adult humans (Anken et al., 2016, 2018; Foxe et al., 2005; Knebel & Murray, 2012; Murray et al., 2002, 2004, 2006; Pegna et al., 2002; Shpaner et al., 2009; Tivadar et al., 2018), we anticipated that responses to the IC and NC conditions would differ, with initial effects peaking at ∼150-170ms post-stimulus onset (reviewed in Murray & Herrmann, 2013). Exemplar VEP waveforms are displayed in **Figure 2** and show that indeed responses were enhanced to the IC vs. NC condition in both groups (see **Figure S2** for GFP waveforms). This was statistically verified by the main effect of Stimulus in the 2×2 rmANOVA on VEP and GFP waveforms and by subsequent paired t-tests within each group. More specifically, analysis of VEP voltage waveforms revealed a significant main effect of Stimulus over three post-stimulus time periods: 138-248ms (all *F’s*_(1,23)_≥ 4.3; *p* ≤ 0.05), 292-329ms (all *F’s*_(1,23)_≥ 4.3*, p ≤* 0.05), and 350-496ms (all *F’s*_(1,23)_≥ 4.3*, p ≤* 0.05). Likewise, analysis of Global Field Power revealed a main effect of Stimulus over similar time periods: 145-203ms (*F*_(1,23)_≥4.78*, p* ≤ 0.04), 254-271ms (*F*_(1,23)_≥4.56*, p* ≤ 0.05), and 343-455ms (*F*_(1,23)_≥4.37*, p* ≤ 0.05). In all cases, the VEPs and GFP were larger/stronger to IC than to NC stimuli. By contrast, there was no evidence of reliable interactions of Group on Stimulus. Topographic clustering provided no indication of any main effect of Stimulus nor interaction involving this factor. Consequently, we focus below on the main effect of Group and interactions involving that factor, as such indicates visual processing differences between myopes and non myopes.

**Figure 2.**
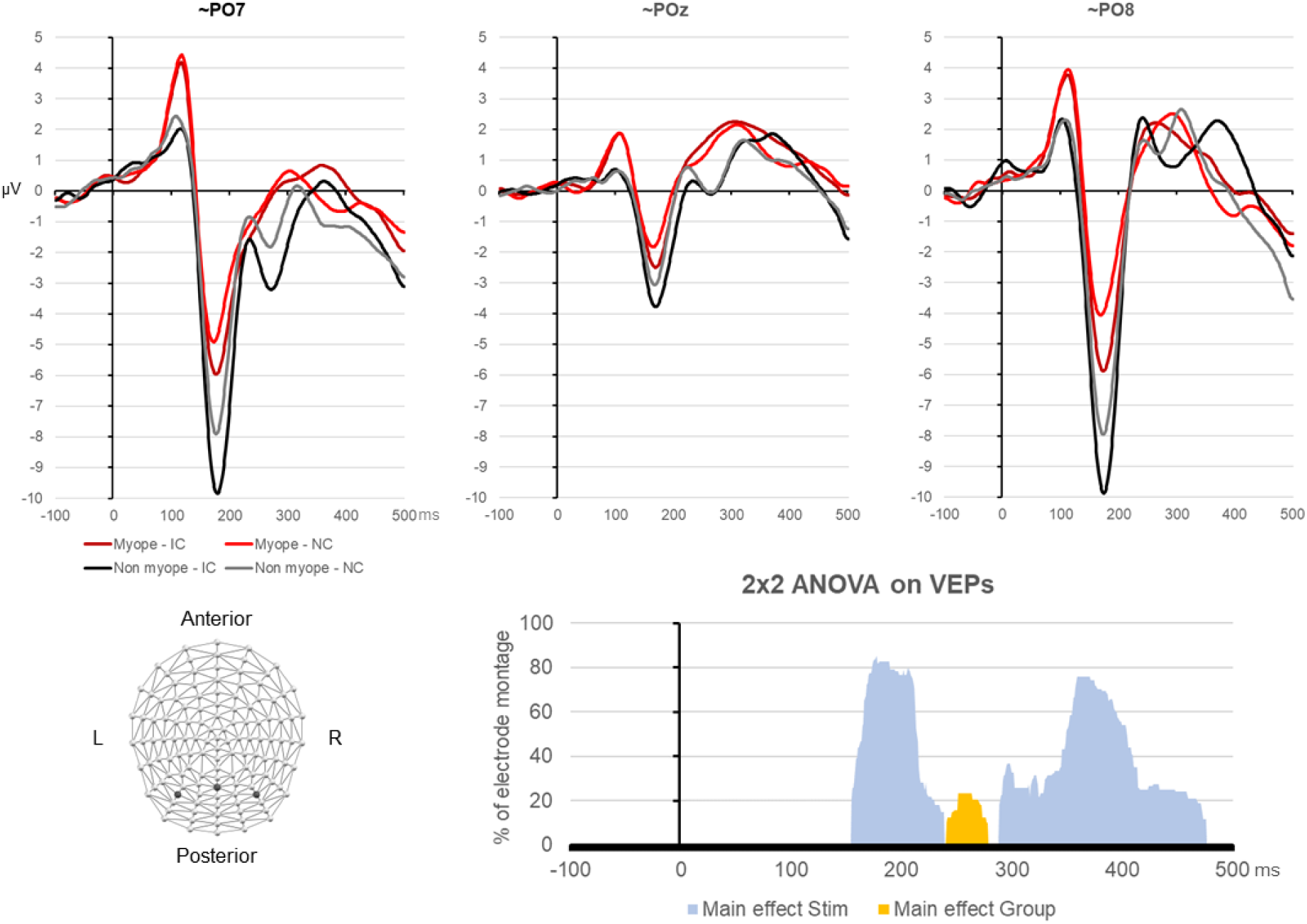
Group-averaged VEP waveforms. The top panels display the group-averaged VEP waveforms at a left parieto-occipital (∼PO7), midline (∼POz) and right parieto-occipital (∼PO8) scalp site in response to IC and NC stimuli (dark and light lines, respectively) for myopes and non myopes (red and black lines, respectively). The bottom left panel illustrates the electrode montage as well as the positions of the 3 electrodes shown in the upper panels. The bottom right panel displays the percentage of electrodes for which the main effect of Stimulus (blue) and the main effect of Group (orange) was significant. Both groups exhibit robust IC sensitivity comparable to prior published works (e.g. Murray and Herrmann, 2013 for review).

#### Group-Wise Differences in Visual Processing

In addition to the main effect of Stimulus, the rmANOVA on VEP waveforms revealed a main effect of Group over the 240-275ms post-stimulus onset period (*F’s* > 4.3; p<0.05; **Figure 2**). The analysis of GFP waveforms provided no evidence for a significant main effect of Group. By contrast, the topographic clustering of the group-averaged data identified a period spanning 218-280ms post-stimulus onset when 3 template maps characterized the data, which we refer to as map2, map3 and map4 (Figure 3a). One of these three template maps appeared to be common to both groups (map2), whereas other template maps (i.e. map3 and map4) appeared to characterize responses from different groups. The single-subject fitting procedure was performed over the 218-280ms post-stimulus period with all three template maps and revealed a significant Group × Template Map interaction (*F*_(1,23)_=6.868; *p*=0.005; *η_p_^2^* =0.384) (Figure 3b). To better understand the bases for this interaction, follow-up ANOVAs were conducted for each template map separately. For map2, there was no evidence for group-wise differences (*F*_(1,23)_=2.582; *p*=0.122; *η_p_^2^*=0.101). By contrast, both map 3 and map 4 exhibited main effects of Group ((*F*_(1,23)_=7.336; *p*=0.013; *η_p_^2^*=0.242) and (*F*_(1,23)_ =9.3; *p*=0.005; *η_p_^2^*=0.290), respectively). More specifically, map3 better characterized responses from myopes than from non myopes (mean±SD = 11.1±2.8ms vs. 0.3±2.9ms), and map4 better characterized responses from non myopes than from myopes (mean±SD = 41.0±5.6ms vs. 17.2±5.4ms). Collectively, these results indicate that responses of myopic individuals result in different topographies of the electric field at the scalp, and by extension different configurations of intracranial sources, than their emmetropic counterparts.

**Figure 3.**
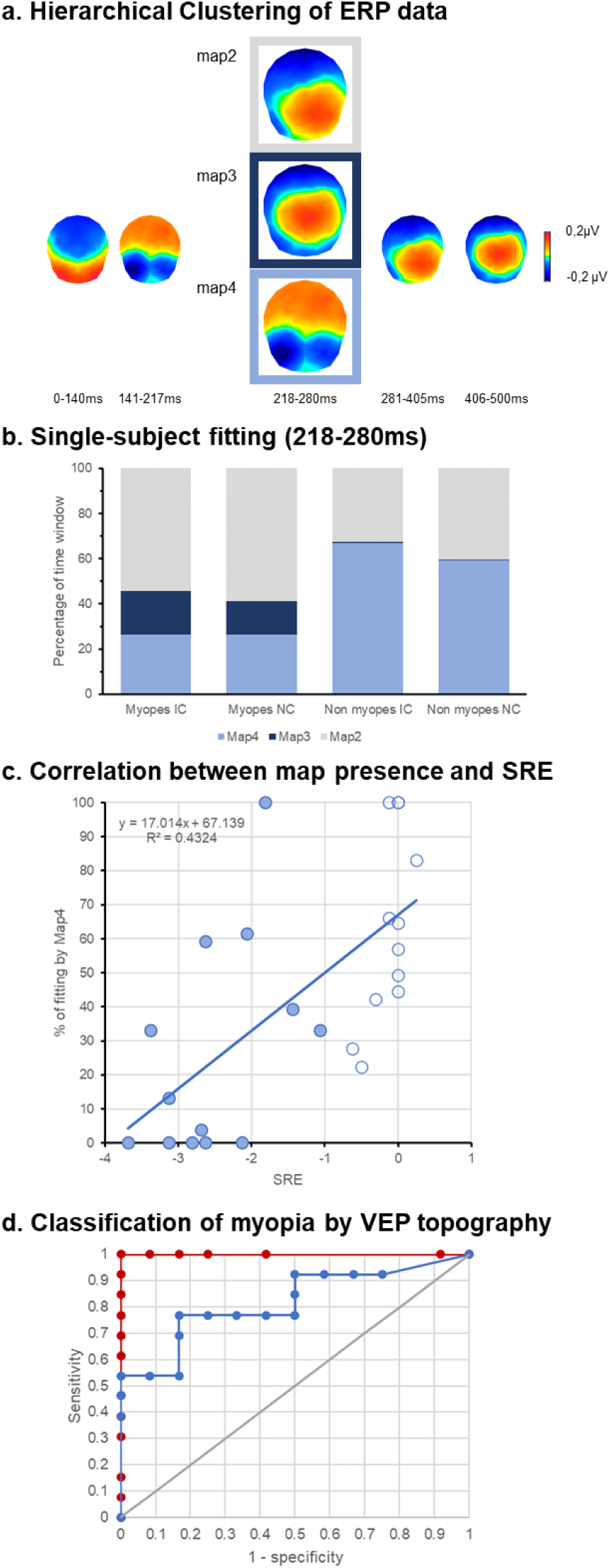
Topographic clustering and single-subject fitting results. a. Topographic clustering identified the 218-280ms time period when different template maps appeared to characterize the group-averaged responses (here denoted map2, map3, and map4). Other time periods were characterized by the same template map across stimulus conditions and groups. b. Single-subject fitting was performed between template maps and revealed that map4 better characterized responses from non myopes, whereas map3 better characterized responses from myopes. There was no evidence that map2 differentially characterized either group. Nor was there evidence for stimulus condition interacting with either factor of map or group. c. The percentage of the 218-280ms time period characterized by map4 positively correlated with SRE, such that the more map4 dominated the VEP response, the less SRE was present. d. The percentage of the 218-280ms time period characterized by map4 could reliably classify an individual as myopic or non myopic. For comparison, we also include the classification based on SRE.

Next, we tested whether the preponderance of map3 and/or map4 was correlated with the extent of refractive error (quantified here by SRE). We first collapsed the fitting data across IC and NC conditions, since there was no evidence of Stimulus condition leading to either a main effect or interaction. There was a significant correlation between the percentage of the 218-280ms period fitted with map4 and SRE (r_(23)_=0.658; p<0.001; Figure 3c). The more the VEP was characterized by map4 during this time period, the higher was the SRE (i.e. less myopic). We furthermore assessed to what extent the VEP topography could reliably classify an individual as myopic, using the area under the ROC curve. For the percentage of the 218-280ms fitted with map4, the accuracy was good (*AUC*=0.817; *SE*=0.087; *p*<0.001 and overall model quality estimated at 0.67). By way of comparison (and as expected), SRE proved to be a perfect predictor (AUC=1.00; *SD*=0.000*, p*<0.0001 and overall model quality estimated at 1.00) (Figure 3d).

Finally, we performed source estimations over the 218-280ms post-stimulus time period to compare groups after collapsing data across stimulus conditions (Figure 5). Robust sources were observed along the medial portion of the occipital pole as well as lateral occipital and inferior temporal cortices bilaterally. The statistical contrast of these source estimations revealed stronger responses from myopes within the medial portion of the occipital pole. Stronger responses from non myopes were observed within inferior frontal and parietal cortices. However, these were diffuse and did not meet our spatial extent criterion.

## Discussion

We provide evidence for general cortical processing differences between non myopes and mild myopes – both of whom were tested under −3D of defocus – that manifest over relatively late post-stimulus periods. That these differences followed from topographic modulations indicates that non myopes and mild myopes engage (partially) distinct networks of brain regions. Our source estimations and analyses thereof localized these differences to the medial portion of the occipital pole, with stronger source activity for myopes than non myopes. Moreover, the predominant topography of the VEP during this time period not only correlated with SRE, but also was an excellent classifier of myopia vs. emmetropia. By contrast, our analyses provided no evidence for differences in visual completion processes; task performance as well as the neural correlates of visual completion were similar in both groups. Nor did we observe differences in visual processing of task-irrelevant dartboard stimuli. Collectively, this pattern of findings supports a model of myopia wherein low-level visual cortices are impacted at relatively late post-stimulus processing stages with task-relevant stimuli, perhaps indicative of altered attentional processes and/or susceptibility to visual fatigue.

Using Kanizsa-type illusory contours, we assessed participants’ abilities to perform visual completion. There was no evidence for group differences in behavior. Rather, performance accuracy was at near ceiling levels, and reaction times were generally faster for IC than NC stimuli in both groups. This pattern suggests that perceptual completion is functionally intact despite blur and myopia. In agreement, McKyton *et al*., (2015) assessed both low-level and mid-level visual functions in sighted and cataract-treated children (see also Hadad *et al.,* 2017). The sighted children were presented with blurred versions of stimuli to emulate the visual acuity of cataract-treated children. The performance of the sighted children presented with blurred stimuli was at near-ceiling levels, like what we observed here. By contrast, performance of cataract-treated children was significantly lower and may reflect a long-term consequence of severe and sustained visual loss during childhood, though longitudinal follow-up of these children remains undone and would be informative regarding the full extent of functional recovery (see also Putzar *et al*., 2007 for similar evidence as well as De Stefani *et al*., 2011 for results of intact IC perception despite retinal scarring in individuals with scotoma from macular degeneration). Similarly, topological perception remains unaffected in corrected-to-normal mild to high myopia (Sun *et al*., 2020). Our results contribute to this topic by showing that adults with mild myopia perform visual completion in a manner indistinguishable from that of non myopes when both groups have the same degree of blur. In agreement, studies of geometric illusions in adults with corrected-to-normal vision in the condition of optical blur also observed that the magnitude of the illusions is not affected by defocus (Ward and Coren, 1976; Coren *et al*., 1978).

Our VEP data provide additional evidence regarding the integrity of visual completion processes despite blur. Both non myopes and myopes exhibited enhanced VEPs to the presence vs. absence of illusory contours; the first phase of which peaked at ∼150-170ms post-stimulus onset (see Figure 2). This enhancement followed from modulations in response strength with no evidence for modulations in response topography. The implication is that visual completion stems from stronger responses within a statistically indistinguishable network of active brain regions. Prior works from our group and others’ have likewise characterized illusory contour processes as unfolding in this manner (e.g. Anken *et al*., 2016; Foxe *et al*., 2005; Knebel and Murray, 2012; Murray *et al*., 2002, 2004, 2006; Murray and Herrmann, 2013; Pegna *et al*., 2002; Shpaner *et al*., 2009; Tivadar *et al*., 2018). Of relevance here are the findings reported in Shpaner et al. (2009) who compared visual completion of illusory contours and salient region stimuli (see also Yoshino *et al*., 2006). Salient region stimuli are comprised of “rounded” versions of the inducers (cf. Figure 1 in Shpaner *et al*. 2009). While not blurred, salient region stimuli have been shown to reduce the strength of perceived completion (e.g. Stanley and Rubin, 2003). What Shpaner *et al*. (2009) demonstrated is that that illusory contour stimuli evoked larger responses at earlier post-stimulus latencies (i.e. ∼150-170ms) than salient region stimuli, which instead evoked larger responses at later post-stimulus latencies. Such results thus differentiate between completion-related effects from the two varieties of stimuli. Our findings here can thus be situated alongside these to support an interpretation wherein robust illusory contour completion occurs during the ∼150-170ms post-stimulus period and despite the presence of −3D blur.

Our principal finding is the presence of general visual processing differences between myopes and non myopes under −3D of defocus during the processing of task-relevant stimuli. Two aspects are particularly noteworthy. On the one hand, the timing of these effects at ∼220ms post-stimulus onset is relatively protracted. On the other hand, group-wise differences followed from topographic rather than strength modulations, indicative of changes in the configuration of the active brain networks. Moreover, source estimations localized differences to medial regions of the occipital pole within primary visual cortices (see Figure 4). The timing of the group-wise differences is roughly 200ms later than reports of cortical response onset, which has been shown to occur at ∼50ms post-stimulus onset in response to high contrast stimuli (e.g. Foxe and Simpson, 2002; Murray *et al.,* 2001, 2002). Moreover, these group-wise differences are subsequent to the initial stages of visual completion, which typically peak at ∼150ms post-stimulus onset as described above. The timing of the present differences at ∼220-280ms instead coincides with reports of periods of more effortful visual completion and visual perception (e.g. Altschuler *et al*., 2012, 2014; Doniger *et al*., 2000, 2001; Foxe *et al*., 2005; Murray *et al*., 2006; Sehatpour *et al*., 2006). For example, Sehatpour et al. (2006) compared brain responses to scrambled and unscrambled drawings of common objects. They observed VEP modulations over the ∼230-400ms consistent with the so-called N_cl_ or negativity for closure that was originally characterized by Doniger *et al*. (2000). Consistent with an interpretation in terms of effortful object recognition, there is evidence that this effect shifts earlier in time, i.e. peaking at ∼150-170ms, when the to-be-recognized object has been identified (Doniger *et al*., 2000). Our results are similar insofar as the group-wise differences reflect changes in the cortical networks recruited by myopes and non myopes to perform visual completion. More specifically, responses from our myopic participants included prominent sources within lateral occipital as well as primary visual cortices, whereas source estimations from emmetropic participants were limited to lateral occipital cortices (**Figure 4**). Although the literature is inconsistent with regard to whether structural alterations in myopia impact gray versus white matter, evidence consistently points to effects within low-level visual cortices along the calcarine sulcus in high myopia (e.g. Huang et al., 2018; Q. Li et al., 2012). In mild myopia, alterations in resting-state cortical dynamics have been described in the optic radiations (Cheng *et al*., 2020a) and outside of visual cortices (Cheng *et al*., 2020b), persisting after refractive surgery (Yu *et al*., 2020).

**Figure 4.**
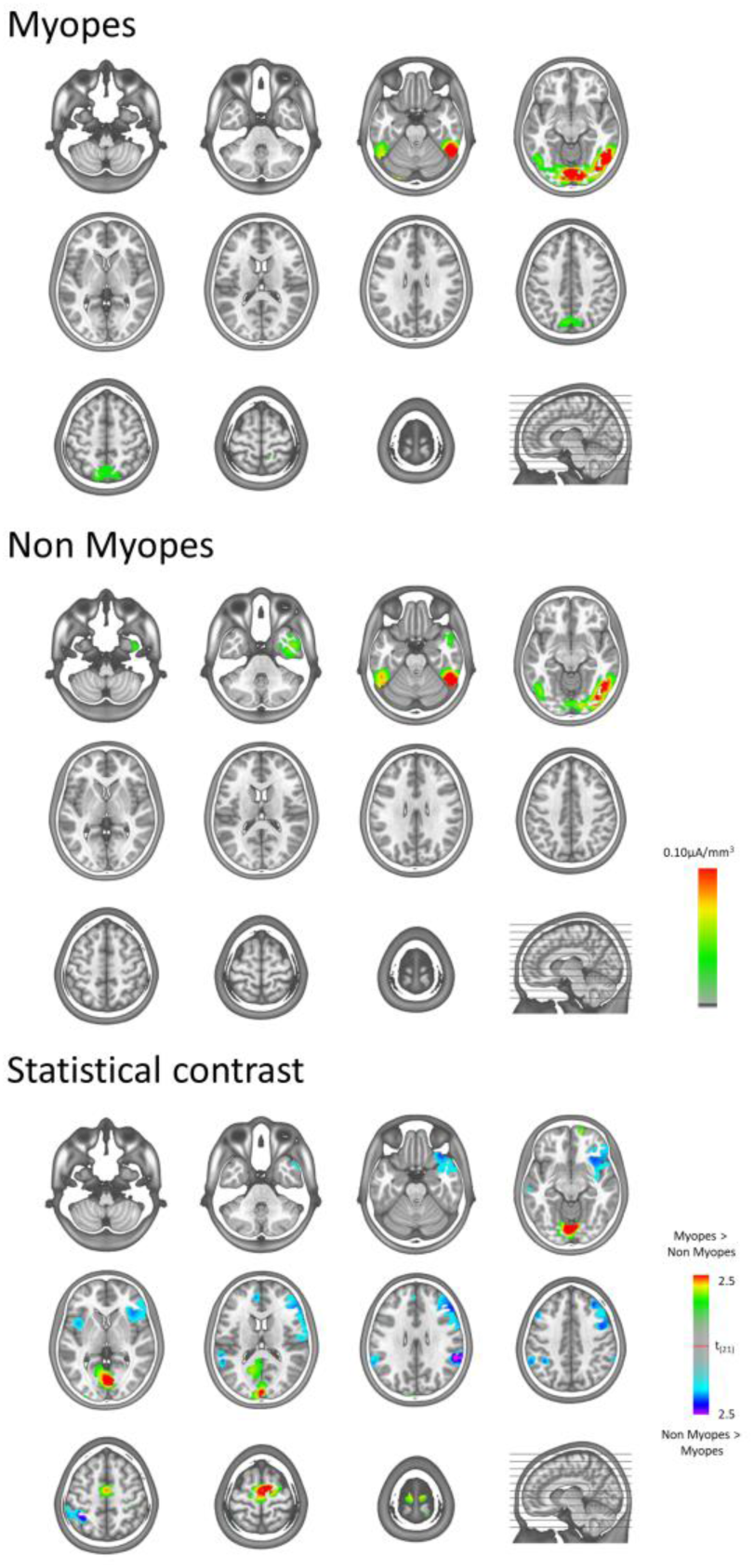
Source modelling data over the 218-280ms post-stimulus period. Group-averaged data are displayed on axial slices (see inset for vertical positions relative to a sagittal slice). Both groups exhibited prominent source activity within the lateral occipital cortices. The group-averaged data from myopes also appeared to include strong source activity within the medial occipital cortices. This was statistically confirmed in the between-groups contrast displayed in the lowermost panel. Differences were considered reliable if they extended spatially over at least 10 contiguous solution points with a contrast as each solution point meeting the p≤0.05 threshold.

Our findings contribute to this line of results by showing that myopic participants more prominently engage lower-level cortices under conditions of equivalent defocus when stimuli are task-relevant. Our analyses of VEPs in response to the task-irrelevant dartboard stimuli provided no evidence for group-wise differences (**Figure S1**). Aligned with these results are reports that myopic individuals retain better contrast sensitivity and reading abilities than emmetropic participants in defocus (Radhakrishnan *et al*., 2002; Poulere *et al*., 2013). It will therefore be of interest to ascertain potential links more finely between activity in low-level cortices and functional abilities in myopia. Likewise, it will be important to determine the full extent to which visual processing differences, like those we report here, have been obfuscated in prior studies of visual completion, such as those cited above, because any myopic participants are typically tested while wearing their corrective lenses and in the absence of defocus. More generally, future research will likely benefit from drawing closer structural - functional relationships regarding effects involving primary visual cortices.

There is now a growing body of work comparing VEPs from myopic participants as well as from emmetropic individuals with defocus transiently induced by defocusing lenses (e.g. Anand *et al*., 2011; Nakamura *et al*., 2014). Most such studies have collected data from a limited number of scalp sites (i.e. often <3) and across a wide range of myopia severity or induced blur. While moderate and severe myopia seems to delay the latency and decrease the amplitude of VEP responses to a pattern-reversing stimulus, no reliable differences were reported between non myopes and mild myopes (i.e. <-3D) (Garg *et al*., 2022). In agreement, we likewise failed to observe differences between non myopes and mild myopes in the processing of the task-irrelevant dartboard stimulus (Figure S1). It should also be noted that Garg et al. (2022) only considered VEP components up to ∼170ms and thus did not examine post-stimulus latencies encompassing the time window here when we observed our group - wise differences. Other studies have found that blur-induced changes on VEP amplitude and latency depend on the stimuli used, with effects occurring for high but not low spatial frequency stimuli (Kordek *et al*., 2022). Specifically, induced blur affected pattern-reversal VEPs, but not motion-onset VEPs. In the case of the present study, the Kanizsa-type stimuli were comprised of a preponderance of lower spatial frequencies, potentially explaining (at least partially) why group-wise effects manifested only at relatively late post-stimulus latencies. Importantly, our results also indicate that such effects are not simply the consequence of blur, as both groups viewed stimuli under identical defocus. Rather, we would contend that the differences we observed here follow from lifetime experience of mild myopia. In agreement with this interpretation is our observation of a strong correlation between the prevailing VEP topography over the 218-280ms post-stimulus period and the extent of refractive error, as measured via SRE. Moreover, the predominance of a given VEP topography was an excellent classifier of an individual as myopic or emmetropic (Figure 3).

In summary, we provide evidence that illusory contour perception and its neural correlates are indistinguishable in myopes and non myopes who are viewing stimuli under equivalent conditions of −3D blur. Despite this similarity, we also characterize general visual processing differences between myopes and non myopes. However, and unlike prior research, we find that these differences manifest at relatively late post-stimulus latencies and stem from topographic differences between groups. Source reconstructions indicated that myopes recruit primary visual cortices during this time, which may reflect a difference in more conceptual phases of object recognition and identification. Finally, the correlation between topographic features of the VEP and measures of refractive error suggests that there may be a direct, longstanding consequence of myopia on visual cortical function.

## Data Availability

All data produced in the present study are available upon reasonable request to the authors.

## Acknowledgements

KS is supported by an MD-PhD fellowship of the Swiss Academy of Medical Sciences (Grant MD-PhD 21/21). MMM is supported by the Swiss National Science Foundation (Grant 169206). We thank Ms. Astrid Minier for their assistance in orthoptic evaluations.

