## Supplementary material for "Cortical Visual Processing Differences in Myopia and Blur": Table S1

Supplementary Table S1: Refraction results in the myopia group.

| *Right Eye* | | | | *Left Eye* | | | | *Avg.* | *Years with* | *Age at First* |
| --- | --- | --- | --- | --- | --- | --- | --- | --- | --- | --- |
| *Sph.* | *Cyl.* | *Axis* | *SRE* | *Sph.* | *Cyl.* | *Axis* | *SRE* | *SRE* | *Correction* | *Diagnosis* |
| -1.50 | -1.00 | 175 | -2.000 | -1.75 | -.75 | 180 | -2.125 | -2.063 | 15-19 | 10-14 |
| -4.00 | -.25 | 70 | -4.125 | -3.25 | .00 | .00 | -3.250 | -3.688 | 10-14 | 10-14 |
| -1.50 | .00 | .00 | -1.500 | -2.00 | -.25 | 120 | -2.125 | -1.813 | 15-19 | 15-19 |
| -2.50 | -1.25 | 10 | -3.125 | -2.50 | -1.25 | 170 | -3.125 | -3.125 | 25-29 | 10-14 |
| -3.25 | .00 | .00 | -3.250 | -3.25 | -.50 | 160 | -3.500 | -3.375 | 15-19 | 10.00-14 |
| -2.75 | -1.00 | 10 | -3.250 | -1.50 | -1.00 | 170 | -2.000 | -2.625 | 5-9 | 15-19 |
| -1.00 | .00 | .00 | -1.000 | -1.00 | -.25 | 110.00 | -1.125 | -1.063 | 10-14 | 10-14 |
| -2.75 | .00 | .00 | -2.750 | -2.50 | -.25 | 155 | -2.625 | -2.688 | 5-9 | 15-19 |
| -2.00 | -.50 | 120 | -2.250 | -3.25 | -.25 | 145 | -3.375 | -2.813 | 10-14 | 10-14 |
| -1.50 | -.25 | 180 | -1.625 | -1.25 | .00 | .00 | -1.250 | -1.438 | 10-14 | 20-24 |
| -2.25 | -.50 | 135 | -2.500 | -2.50 | -.50 | 180 | -2.750 | -2.625 | 25-29 | 10-14 |
| -3.25 | .00 | .00 | -3.250 | -3.00 | .00 | .00 | -3.000 | -3.125 | 5-9 | 15-19 |
| -1.00 | -1.25 | 75 | -1.625 |  |  |  | -2.625 | -2.125 | 10-14 | 10-14 |
