## Supplementary material for "Cortical Visual Processing Differences in Myopia and Blur": Table S2

Supplementary Table S2: Refraction results in the emmetropia group.

| *Right Eye* | | | | *Left Eye* | | | | *Avg.* |
| --- | --- | --- | --- | --- | --- | --- | --- | --- |
| *Sph.* | *Cyl.* | *Axis* | *SRE* | *Sph.* | *Cyl.* | *Axis* | *SRE* | *SRE* |
| .25 | .00 | .00 | .250 | .25 | .00 | .00 | .250 | .250 |
| -.25 | -.25 | 160.00 | -.375 | -.25 | .00 | .00 | -.250 | -.313 |
| .00 | .00 | .00 | .000 | .25 | -.50 | 180.00 | .000 | .000 |
| -.25 | -.25 | 120.00 | -.375 | -.75 | -.25 | 90.00 | -.875 | -.625 |
| -.25 | -.50 | 110.00 | -.500 | -.25 | -.50 | 40.00 | -.500 | -.500 |
| .00 | .00 | .00 | .000 | .00 | .00 | .00 | .000 | .000 |
| .00 | .00 | .00 | .000 | .00 | .00 | .00 | .000 | .000 |
| .00 | .00 | .00 | .000 | .00 | .00 | .00 | .000 | .000 |
| .00 | .00 | .00 | .000 | .00 | .00 | .00 | .000 | .000 |
| -.25 | .00 | .00 | -.250 | .00 | .00 | .00 | .000 | -.125 |
| -.25 | .00 | .00 | -.250 | .00 | .00 | .00 | .000 | -.125 |
| .00 | .00 | .00 | .000 | .00 | .00 | .00 | .000 | .000 |
