## Supplementary material for "Cortical Visual Processing Differences in Myopia and Blur": Figure S1

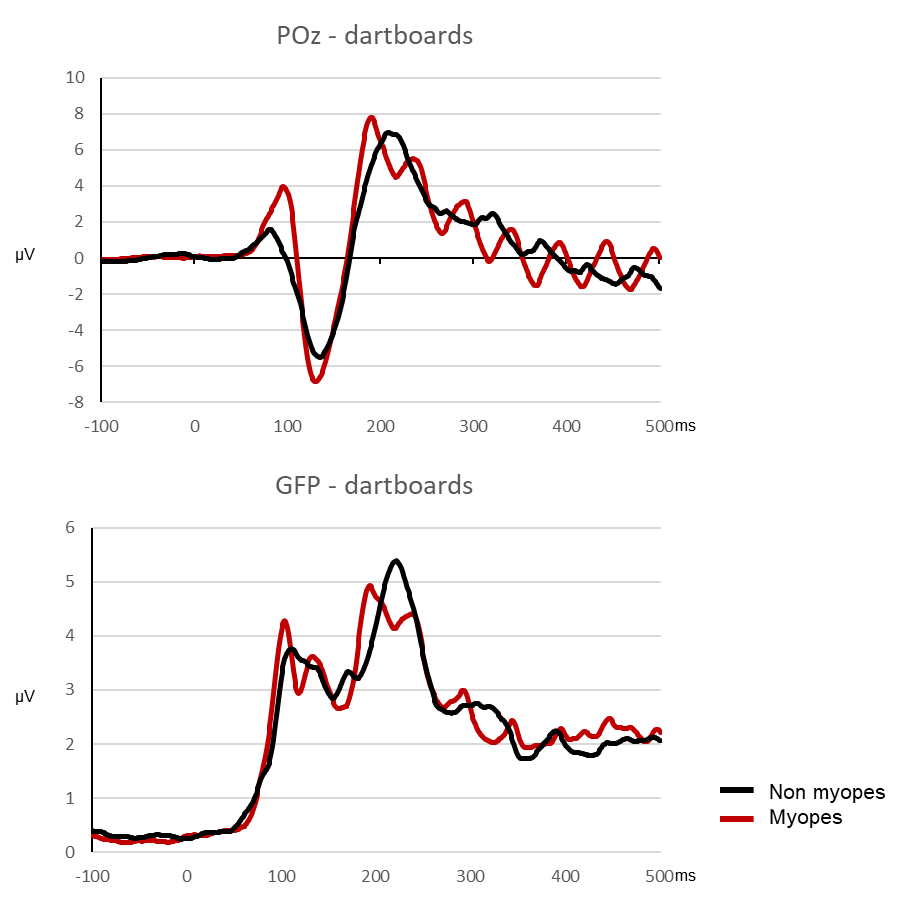

**Figure S1.** The upper panel displays group-averaged VEPs from an exemplar midline parieto-occipital electrode (POz) in response to the task-irrelevant dartboard stimulus. The lower panel displays the group-averaged Global Field Power (GFP). The shaded area around the traces display the s.e.m. The black lines refer to data from non-myopes, and the red lines from myopes. There was no evidence of reliable differences either at the VEP waveform or GFP level.
