## Supplementary material for "Cortical Visual Processing Differences in Myopia and Blur": Figure S2

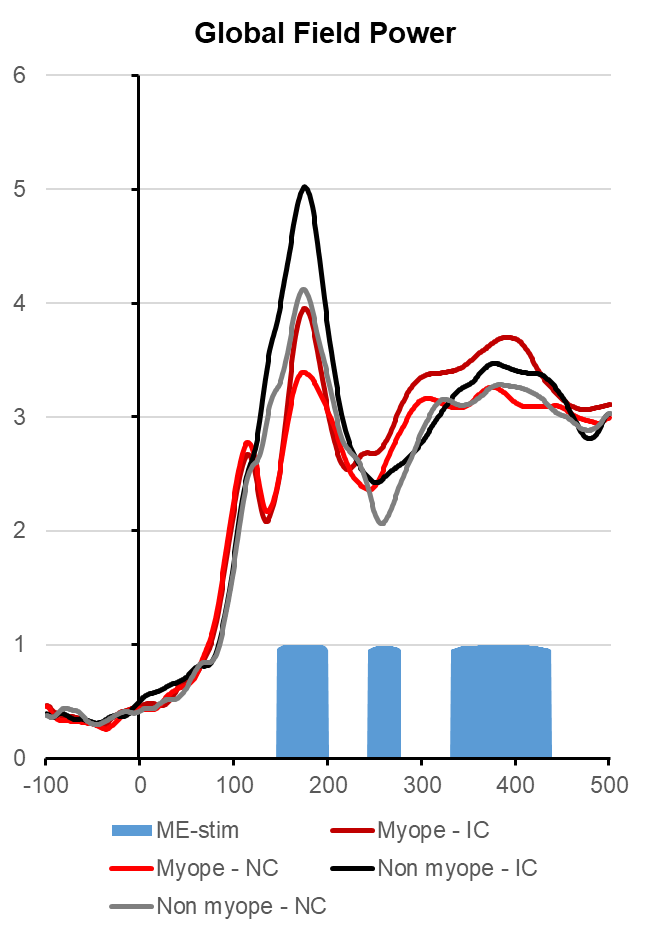

**Figure S2.** Group-averaged GFP waveforms from myopes and non-myopes in response to the IC and NC stimulus condition. The blue area plot shows time periods of a significant main effect of stimulus. No other main effect nor interaction was statistically reliable.
